# An Early Assessment of Curfew and Second COVID-19 Lock-down on Virus Propagation in France

**DOI:** 10.1101/2020.11.11.20230243

**Authors:** Christelle Baunez, Mickael Degoulet, Stéphane Luchini, Patrick A. Pintus, Miriam Teschl

## Abstract

This note provides an early assessment of the reinforced measures to curb the COVID-19 pandemic in France, which include a curfew of selected areas and culminate in a second COVID-19-related lock-down that started on October 30, 2020 and is still ongoing. We analyse the change in virus propagation across age groups and across départements using an acceleration index introduced in Baunez et al. (2020). We find that while the pandemic is still in the acceleration regime, acceleration decreased notably with curfew measures and this more rapidly so for the more vulnerable population group, that is, for people older than 60. Acceleration continued to decline under lock-down, but more so for the active population under 60 than for those above 60. For the youngest population aged 0 to 19, curfew measures did not reduce acceleration but lock-down does. This suggests that if health policies aim at protecting the elderly population generally more at risk to suffer severe consequences from COVID-19, curfew measures may be effective enough. However, looking at the departmental map of France, we find that curfews have not necessarily been imposed in départements where acceleration was the largest.

**JEL Classification Numbers:** I18; H12

## 1 Introduction

In this note, we provide an early assessment of reinforced measures to dampen the spread of COVID-19 in France. These measures where introduced in three waves, each with one week interval. First, from October 17 onwards, eight major cities and their adjacent surroundings called “métropoles”, plus the whole of “Île de France” which includes Paris, all together corresponding to 16 “départements” (roughly counties) were put under curfew from 9pm to 6am.^1^ The following week, 38 départements were further added, so that a total of 54 out of 96 départements of metropolitan France were declared under mandatory curfew from October 23 onwards. Finally, starting on October 30, a nation-wide lock-down was implemented and is still ongoing. These three waves of health policy measures to combat the COVID-19 pandemic provide an excellent data-set to compare. In the following, we refer to those three weeks respectively “curfew 1”, “curfew 2” and “lock-down”.

We first focus on virus propagation of two age groups, below and above 60 years, and across départements. These two age groups correspond roughly to the population which is more (above 60 years old) and less (59 years and younger) vulnerable in terms of hospitalisation and mortality (e.g. Promislow [4]). We also single out in particular the age group of young people aged 19 and less as they continue to go to school even though curfew and lock-down measures were put in place. To make this assessment, we update the acceleration index introduced in Baunez et al. [1]. This index tracks in real-time the propagation of COVID-19 and can do so at very granular levels. It is a scale-free measure of the speed at which tests detect positive cases. This acceleration index aggregates information provided by the daily and average positivity rates, as computed from daily numbers of tests and confirmed cases. This means that the index tracks in real-time the acceleration and deceleration of the the virus and can thus serve to visualise rapidly whether any health policy measures have their desired effects or not.

The main results are that the curfew on a larger number of départements in week 2 seems almost as effective in dampening the acceleration among the age group 60+ who are particularly at risk of severe COVID-19 consequences as lock-down measures. Lock-down measures do have quicker effects on both age-groups and especially on the population 59 and younger. Curfew measures do not have any effects on reducing acceleration among the youngest population aged 0 to 19, but lock-down does. However, looking at the departemental map of France, we find that curfews have not necessarily been imposed on départements which did have the greatest acceleration. It is therefore difficult to judge unambiguously which of the policy measures was the most effective one, other than to say that lock-down measures have a quicker impact on the whole population, but very possibly with much higher economic and social costs than curfew measures. Given that the population aged 60 and more is notoriously more at risk of severe COVID-19 consequences, our data analysis may suggest that curfew measures are enough to reduce the viral spread among this group.

In the following we present the effects of curfew and lock-down measures by looking in particular at the two age-groups 60 years and more, as well as 59 years and younger (section 2.1). In section 2.2, we single out the effect of reinforced COVID-19 measures on the youngest generation aged 0 to 19. Section 2.3 presents the analysis per départements over the three waves of COVID-19 measures for the two age groups.

## 2 The Effects of Curfew and Second Lock-down in France: an Early Assessment using the Acceleration Index

As stressed in Baunez et al. [1], accurate information about the dynamics of a pandemic rests on both the number of cases and the number of tests, and that the former cannot be properly understood without the latter. In the aforementioned paper, we address this pressing issue and provide an acceleration index that relates to the daily and average positivity rates, in the following way. From data on the cumulated numbers of confirmed cases and tests, up to final date *T*, and if we denote the acceleration index at end date *ε*_*T*_, the following decomposition holds:

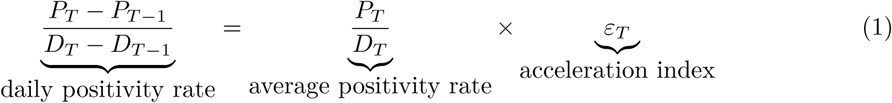

where *P*_*T*_ and *D*_*T*_ denote respectively the cumulated numbers of positives/cases and diagnosis/tests at date *T*, and similarly for date *T* − 1. Therefore, the decomposition in equation (1) shows that the acceleration index is defined as the ratio of the daily positivity rate to the average positivity rate, that is, it is essentially an elasticity.

### 2.1 Effects on People Aged 60 and Older Relative to All Other People

In Figure 1 we report in panel (*a*) the daily and average positivity rates, while in panel (*b*) is depicted the acceleration index, for the age groups of people aged 59 and less (solid lines), on the one hand, and of people older than 60, on the other (dashed lines).

**Figure 1:**
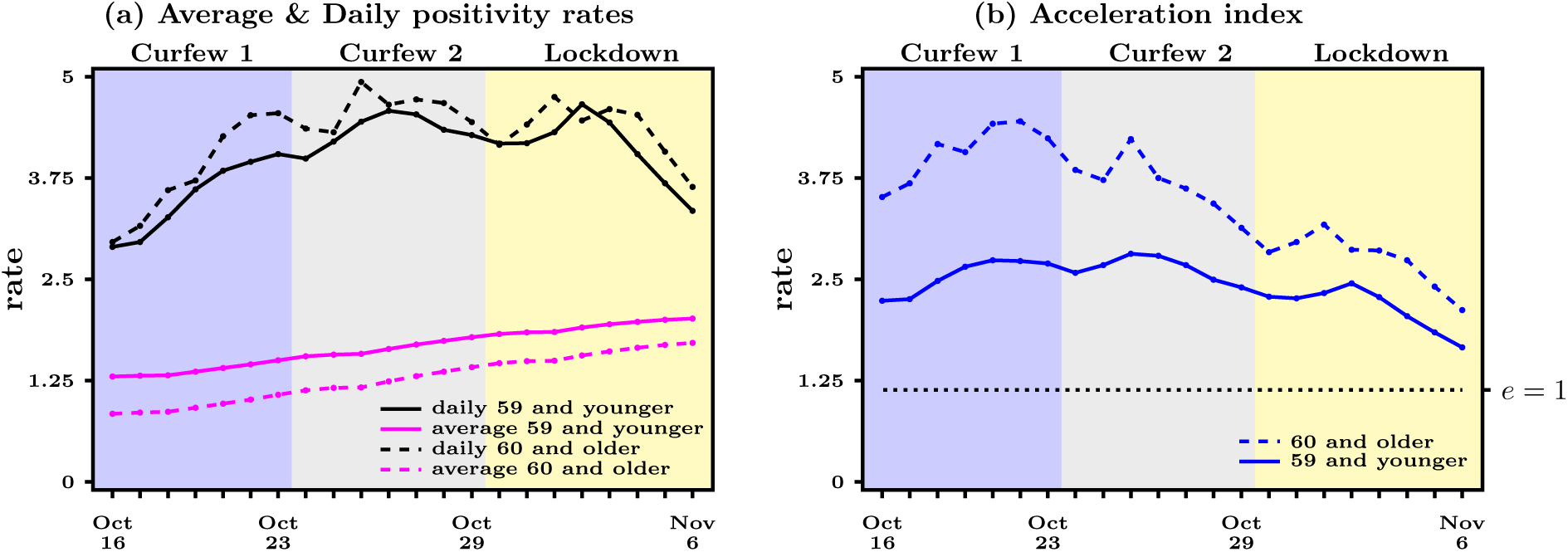
Main acceleration indicators for different age groups - daily positivity rate (daily cases to daily tests ratio, black line) and average positivity rate (daily cumulated cases to daily cumulated tests ratio, purple line) over time in left panel; acceleration index (ratio of daily positivity rate to average positivity rate, blue line) over time in right panel, October 16 - November 6, 2020. Dashed horizontal line in right panel represents when acceleration index equals 1. Data source: Agence Santé Publique France

What is particularly striking in panel (*b*) of Figure 1 is that the acceleration index still increases during curfew 1 (Oct 17 - Oct 23) for both age groups when only 16 departments were under curfew, but started to decline during curfew 2 (Oct 24 - Oct 29), when the curfew was extended to 54 départements in France. Acceleration continues to decline markedly during the first week of lock-down (Oct 30 - Nov 6) for both age groups. The decline over the two last weeks is particularly true for the age group 60 and older. While their index was at 3.69 on Oct 17, it raised to 3.72 during curfew 1 on Oct 24 and then declined to 2.83 during curfew 2 and to 2.12 at the end of the first week of the lock-down. The percentage change of the acceleration index during curfew 2 was -23.9% and during the first week of the lock-down it was -25.2%. Looking at these numbers, it seems that curfew 2 had quite a considerable effect in terms of reducing acceleration for the more vulnerable group, with the lock-down having only a slightly better impact. The acceleration index for the age group 59 and younger however decreased during curfew 2 by only -14.5%, but decreased more rapidly by -27.3% during the first week of the lock-down. Hence it seems that while the curfew works for the age group 60+, it is especially the lock-down that affects the mostly active age group of 59 and younger. This appears intuitive as more active people are working from home or, in the worst case, cannot further engage in their professional activities and/or lose their jobs during lock-down and are thus supposedly less exposed to social contacts and transmission channels.

Looking at panel (*a*) in Figure 1, we see that the decline of the acceleration index for both age groups combines (*i*) a steadily increasing average positivity rate, which goes in the direction of decreasing the acceleration index, with (*ii*) a reversal of the daily positivity rate, which increases during curfew 1 and 2 but goes down during the first week of lock-down. This indicates that looking at daily or average positivity rate only does not deliver the same information as the acceleration index, or, if the trend persists, only at a much later stage.

All of the above suggests that the curfew has been rather effective, especially for the elderly population generally more at risk of severe consequences of COVID-19, including death. The momentum gained in terms of reducing virus circulation seems to be reinforced by the lock-down, affecting the population more generally, but at higher social and economic costs that are widely discussed elsewhere (e.g. Blanchard et al. 2020 [2]). It should also be noted that according to governmental data, a vast majority of the 60+ generation are admitted in hospitals and intensive care in French hospitals these days.^2^ If the aim of the reinforced health policy measures is to reduce the viral spreading among the group of people most at risk of hospitalisation and death, then by looking at the data as we have done above, the question may be raised whether a curfew would not be enough.

### 2.2 Effects on People Aged 19 and Younger

In Baunez et al. [1], we show that the acceleration index increases rapidly for children aged 0 to 9 since early September. Before September, acceleration for that age group declined to reach the acceleration/deceleration threshold, that is an acceleration index of about 1, by the end of August (while the national average was still indicating acceleration). But since then, schools opened again and the acceleration increased to reach levels higher than national average. However, we did not observe such a sharp acceleration for the age group from 10 to 19 since schools opened again. Acceleration increased as well but stayed below national average. We therefore look more specifically at the impact of the reinforced COVID-19 measures on the acceleration of the younger generation up to 19 years old, which is a widely discussed topic in academic and media circles. In Figure 2, we see that for both age groups, 0-9 and 10-19, curfews 1 and 2 do not stop the acceleration index from increasing, while lock-down does. This is to be expected since curfew measures do not affect most of the younger people’s activities. However, even though schools remained open even during lock-down, an effect is visible, which could be linked to decreased physical and social activities outside school. The decrease is more marked for the 10 to 19 years old, than for the youngest children, who are possibly at an age where they engage in fewer social activities outside school than teenagers. However, the youngest children still face the greater acceleration. The acceleration index for children aged 0-9 was about 3.94 at the beginning of lock-down on October 30, but fell during the first week of lock-down by -19.5% to 3.17 on November 6. Their acceleration is thus well above the acceleration of people aged 60+, whose index was higher than of the whole group of people aged 59 and less. The acceleration index for teenagers aged 10-19 equaled about 2.72 on October 30 and fell by -28.5% to 1.95. It has to be stressed however that to the extent that children are in general at a very low risk of hospitalisation,^3^ acceleration among younger people are only worrying if they are in contact with the people most at risk of suffering from COVID-19 consequences. It seems therefore important to separate children from the population most at risk of having severe consequences from COVID-19. Whether a lock-down is the right measure to do so seems to be a valid question. Consequently, it may also be questioned whether a reinforced lock-down as some demand, which would include school closures, would be the right policy to take, in particular if the current trend of reduced acceleration among children persists.

**Figure 2:**
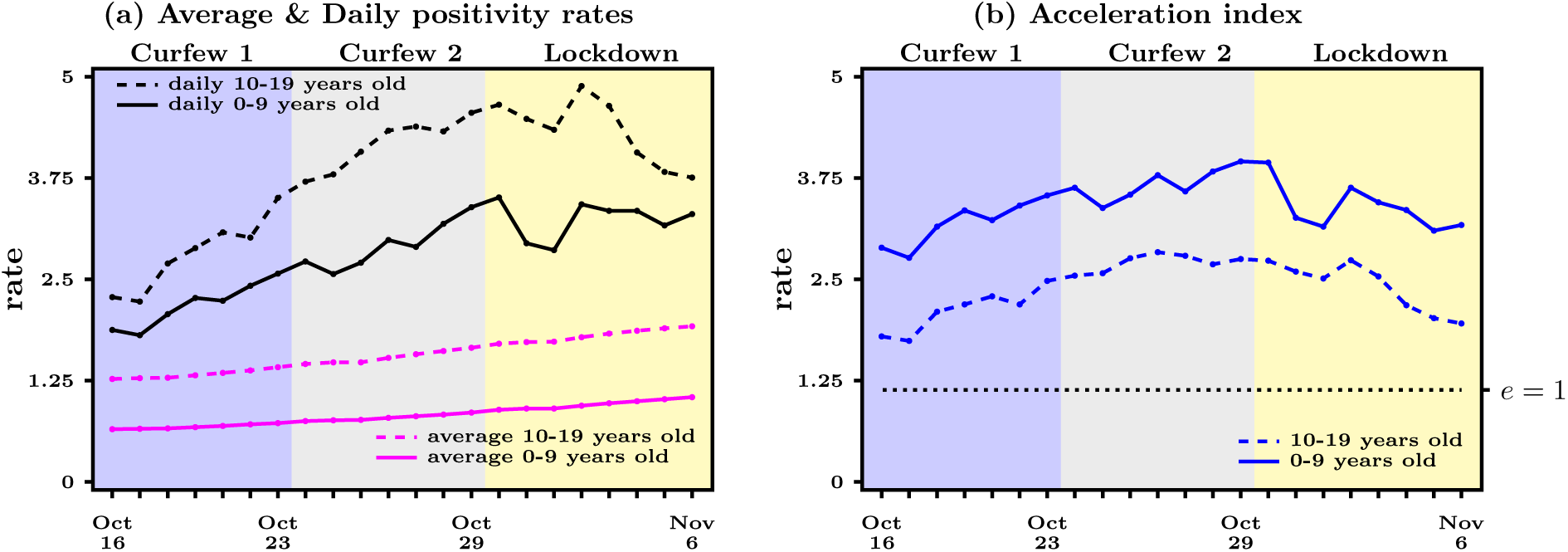
Main acceleration indicators for young age groups - daily positivity rate (daily cases to daily tests ratio, black line) and average positivity rate (daily cumulated cases to daily cumulated tests ratio, purple line) over time in left panel; acceleration index (ratio of daily positivity rate to average positivity rate, blue line) over time in right panel, October 16 - November 6, 2020. Dashed horizontal line in right panel represents when acceleration index equals 1. Data source: Agence Santé Publique France

### 2.3 Effects across Départements

In Figure 3, we report the weekly averages of the acceleration index across départements, for the age groups of people aged 59 and less (right maps), on the one hand, and of people older than 60, on the other (left maps).

**Figure 3:**
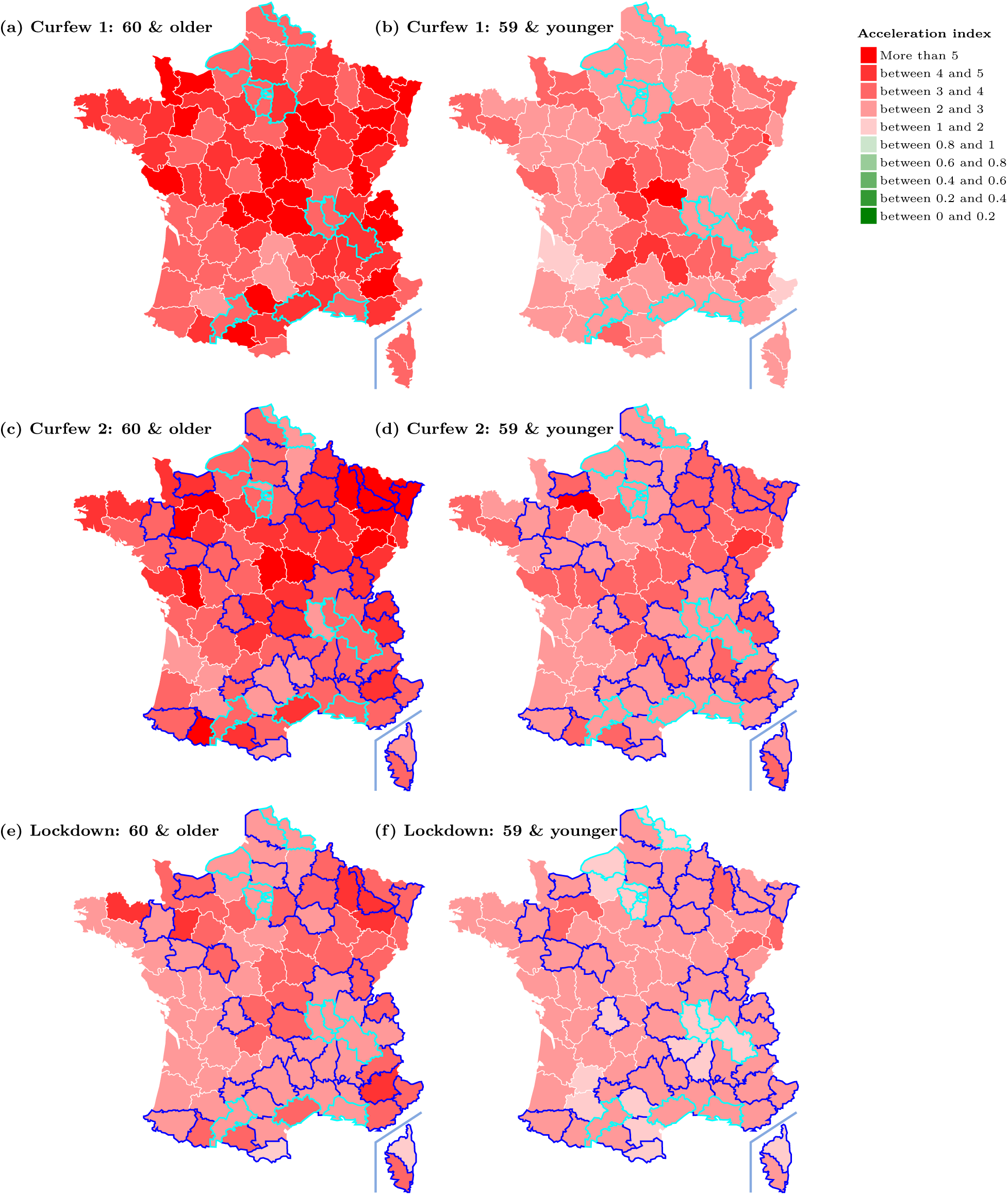
Acceleration index for French départements for both age groups (weekly averages for the 3 consecutive weeks also shown in Figure 1 and 2). Contours in light (dark) blue depict départements subject to both Curfew 1 and Curfew 2 (Curfew 2 only). Data source: Agence Santé Publique France

What we can clearly see in Figure 3 is first, that even across départements, acceleration is generally higher for the age group 60+ than for those aged 59 and younger. Second, curfew 1 and 2 have not necessarily been imposed on those départements that experienced the strongest acceleration. Rather, curfew 1 in particular, but also curfew 2 have been imposed on larger metropolitan areas with larger absolute numbers of positive cases. The official public health decision regarding which départements to put under curfew were based on the following governmental criteria: (*i*) more than 250 positive cases per 100 000 inhabitants, (*ii*) more than 100 cases per 100 000 elderly inhabitants; and (*iii*) the occupation rate of intensive care beds starts to exceed 30% with a tendency to increase further in the following weeks. As we show in Baunez et al. 2020 ([1]), the current testing strategy in place closely follows population numbers. It is then not surprising to find more cases when more tests are conducted in more populated areas. This explains why the selected dpártements were put under curfew rather than those where the acceleration is greatest. However, as we also show in Baunez et al. [1], more positive cases are not necessarily equivalent to greater acceleration. It may therefore be asked whether the governmental selection criteria were the right ones in the first place. Our guess is that curfew measures would have had an even stronger impact if they had been imposed on départements with the greatest acceleration from the very beginning.

From Figure 3, we also learn that none of the départments has switched from an acceleration regime to the deceleration regime. The whole map remains in different shades of red, but none of the départments turns green, which would indicate deceleration. The first two maps on the top have the 16 départments under curfew 1 encircled in light blue. The next two maps have the additional 38 départments under curfew 2 marked in dark blue. The last two maps represent the acceleration index during the first week of the lock-down for the two age-groups. We again observe that the reduction in the acceleration index for people older than 60 is more pronounced across the départments on which a curfew was imposed than for the people aged 59 or younger. The trend for a reduced acceleration continues during the first week of the lock-down for both age groups across France.

To sharpen the conclusions from the visual inspection of Figure 3, we focus on the development of the acceleration index over the three waves of COVID-19 policy measures of the two age groups across départments in three different scenarios or “treatments”. These “treatments” are: départements under curfew 1 and more (*curfew 1+*), départements under curfew 2 and more (*curfew 2+*), and départements under lock-down only (*lock-down only*). In panel (*a*) of Figure 4, we report the acceleration index for the age group of people 59 and younger, while panel (*b*) takes care of the people 60 and older.

**Figure 4:**
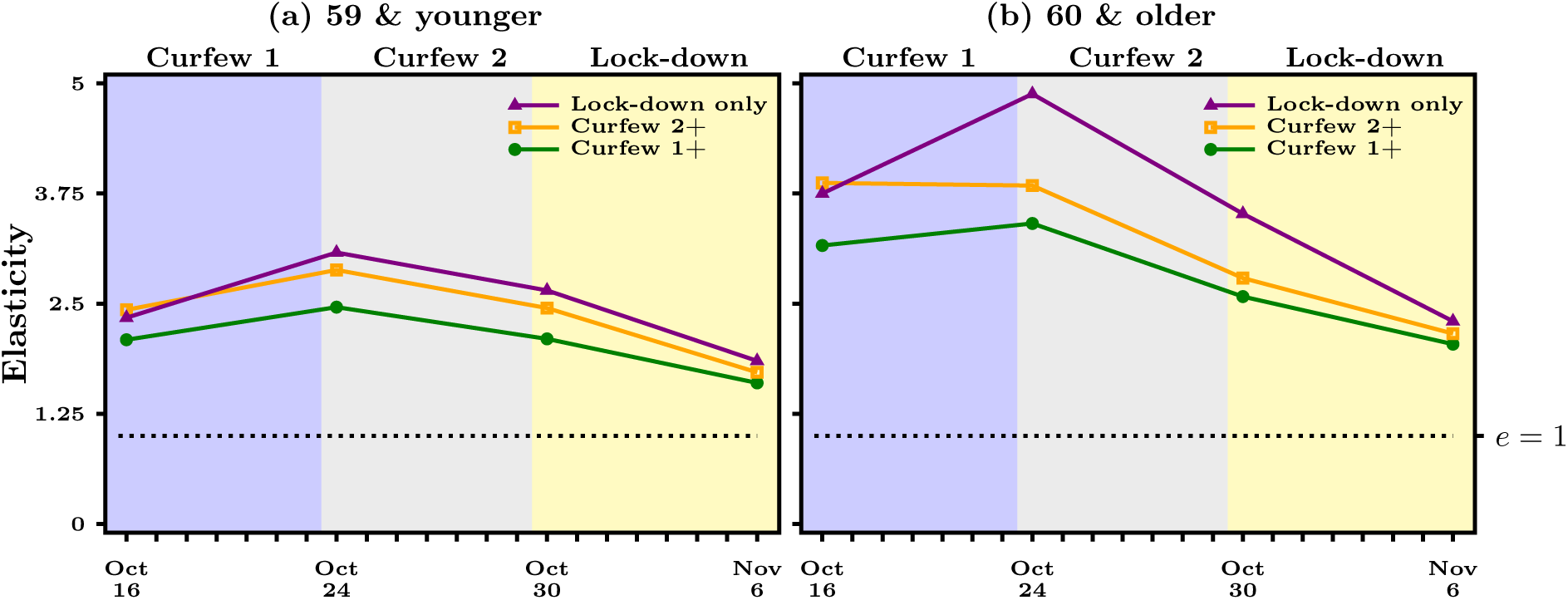
Acceleration index for French départements in different treatment groups (Lock-down only, curfew 2+ for second week of curfew followed by lock-down, curfew 1+ for two weeks of curfew followed by lock-down) for both age groups (weekly averages for the 3 consecutive weeks also shown in Figure 1). Dashed horizontal line in right panel represents when acceleration index equals 1. Data source: Agence Santé Publique France

Figure 4 confirms that the dpártements in “treatments” *curfew 1+* and *curfew 2+* have not been the ones where virus circulation was accelerating the most. This is especially the case for the 60+ who live in the départements of the *lock-down only* and the départements of the *curfew 2* group. A similar conclusion holds for the people aged 59 and younger. As a result, it comes as no surprise that the ongoing lock-down has a stronger effect on reducing acceleration for the 60+ who live in the *lock-down only* départements. What we also see is that all three “treatments” achieve by and about the same acceleration index by Nov 6. Unfortunately, looking at this graph indicates that nothing more general can be said about which is the most efficient policy measure to the extent that, as we have said repeatedly above, the selection criteria for curfew has not been based on the acceleration of the COVID-19 spread. What is remarkable though is that curfew measures achieve by and large similar effects but over a longer period of time than a lock-down. Said differently, a general lock-down has, unexpectedly, quicker effects than curfew only measures. However, the collateral economic and social costs may well be higher with lock-down measures than with curfew measures.

## 3 Conclusion

This note provides an early assessment of the curfew and lock-down measures that have been adopted to curb the COVID-19 pandemic in France. We analyse the change in virus propagation across age groups and across départements using an acceleration index introduced in Baunez et al. (2020) [1]. We find that while the pandemic is still in the acceleration regime, acceleration decreased notably with curfew measures and this more rapidly so for the more vulnerable population group, that is, for people older than 60. Acceleration continued to decline under lock-down, in particular for the active population aged 59 and younger in comparison to curfew measures. For the youngest population aged 0 to 19, curfew measures did not reduce acceleration but lock-down does. This suggests that if health policies aim at protecting the elderly population generally more at risk to suffer severe consequences from COVID-19, curfew measures may be effective enough. Obviously, common sense would then also suggest that such measures should be accompanied with increased sanitary and social protection of the most vulnerable in this current pandemic situation as well as compensation for those who suffer from any economic losses among the general population due to the curfew. Unfortunately though, looking at the departmental map of France, we find that curfews have not necessarily been imposed in départements where acceleration was the largest, which makes conclusive comparisons of the effectiveness of COVID-19 measures harder to do. Our suggestion however is that curfew and lock-down measures would have gained from following the logic of our acceleration indicator.

In France and in other European countries, concerns about both the efficacy and the economic and social costs of a general lock-down, even in its “light form” have been raised extensively. Part of the theoretical literature has suggested that government might consider age-specific strategies that, in particular, allow to protect the most vulnerable citizens while allowing virus circulation to build up collective immunity. Gollier (2020) [3], among others, make such an argument more formal within the setting of Susceptible-Infected-Recovered models at the country level.

The present contribution reinforces this conclusion, but by proposing an analysis of the existing data in real-time, based on our acceleration index. This index is particularly relevant to help assessing what the effects of the ongoing lock-down are, as it is applicable to data that are disaggregated geographically and also in the cross-section in terms of age-groups. This is a major advantage with respect to existing models that are hardly amenable to very granular levels and are subject to considerable parameter-uncertainty, especially at times when a novel pathogen emerges, as in the case of SARS-Cov2.

Our analysis clearly shows that the COVID-19 pandemic spreads differently within different age-groups and across space. It has become clear over the last few months since this novel pathogen emerged earlier this year that the age-group 60+ is the main group at risk of sever consequences due to COVID-19. Among the people currently in hospital, a vast majority of them are 60 years old and more. The same applies to dying from COVID-19. Other age-groups do not, in general, face the same severity even though they are currently exposed to by and large similar acceleration levels. Given this information and the adaptability of our indicator to local and age-specific but potentially also other characteristics, it can be used to guide health policy measures with the very specific aim to bring, in real time, the pandemic under control especially for the population most at risk and to keep them safe without imposing disproportional social and economic costs on the other population groups.

## Data Availability

Data publicly available at Agence Santé Publique France

## Footnotes

1 A caveat is in order to indicate that the eight métropoles are not exactly corresponding to the départements in which they are. However, in the following, we consider métropoles to correspond to départements and therefore refer to 16 départements for the first curfew, one for each of the eight métropoles and the eight départements to which corresponds “Île de France”.

2 Hospital data is publicly available via https://geodes.santepubliquefrance.fr/, which gives the number of persons in hospital and intensive care by age group on a daily basis, as well as mortality rates for all age groups. On November 11, 31,477 are in hospital with COVID-19 and 84.6% of them are 60 years and older. Covid related deaths are also concentrated in the older age group: 93.8% of the deceased where 60 years old or older on November 10.

3 Hospital data at https://geodes.santepubliquefrance.fr/ show that by November 11, 106 children aged 0-9 were in hospital which represents 0.3% of all hospitalised people. That same day, 71 teenagers were in hospital, which represented 0.2% of all hospitalised people.

